# Hyperbaric oxygen therapy improves clinical symptoms and functional capacity and restores thalamic connectivity in ME/CFS

**DOI:** 10.1101/2025.10.29.25339096

**Authors:** Laura Kim, Guido Cammà, Claudia Kedor Peters, Maron Mantwill, Oliver Müller, Nadège Leprêtre, Cornelia Heindrich, Rebekka Rust, Moritz Krill, Tim J. Hartung, Lukas G. Reeß, Stephan Krohn, Christian von Heymann, Kirsten Wittke, Carsten Finke, Carmen Scheibenbogen

## Abstract

Background: Myalgic encephalomyelitis/chronic fatigue syndrome (ME/CFS) is a debilitating disorder characterized by profound fatigue, cognitive impairment, autonomic dysfunction, and exertional intolerance with strongly impaired physical functioning. Hyperbaric oxygen therapy (HBOT) has been proposed as a potential treatment, but its effects on ME/CFS patients remain largely unexplored. This study aimed to evaluate the effectiveness and feasibility of HBOT in ME/CFS patients and to investigate its effects on functional brain changes.

Methods: 30 ME/CFS patients (mean age: 42·3 ± 11·7 years; seven males, 23 females) received 40 HBOT sessions each. Outcomes were assessed at baseline, during treatment, and four weeks post-treatment. The primary outcome was change in the Physical Functioning subscale of the Short Form-36 Health Survey (SF-36 PF). Secondary outcomes included severity of core symptoms assessed via questionnaires, exercise capacity, handgrip strength, cognitive per-formance, orthostatic intolerance, and brain MRI (volumetry and functional connectivity, (FC)). Thirty age- and sex-matched healthy controls (HCs) (mean age: 42·3 ± 11·3 years; seven males, 23 females) were included for MRI comparison.

Findings: In the linear mixed model, SF-36 PF significantly improved during HBOT compared with baseline (g = 0.71, p = 0.006). SF36 pain (p = 0·002, g = 0·79) and CFQ fatigue showed clinically meaningful reductions (p < 0·001, g = - 0·87) during HBOT. Exercise capacity (g = 0·66), muscle strength (g = 0·40), and information processing speed (g = 0·52), all improved significantly after HBOT compared to baseline (all p < 0·05). Treatment adherence was high, and tolerability was favorable, with no major side effects reported. Functional MRI analyses revealed increased thalamic FC in ME/CFS patients compared to HCs in bilateral sensorimotor (p < 0·001, t = 5·65, FDR-corrected) and visuo-occipital regions (p < 0·001, t = 5·4, FDR-corrected) at baseline. After HBOT, thalamic hyperconnectivity normalized. Responders (defined as a ≥ 10 point increase in SF-36 PF) showed greater reductions in thalamic hyperconnectivity than non-responders (p < 0·001, t = - 4·34 to -5·18, FDR-corrected).

Interpretation: HBOT was well-tolerated and was associated with significant improvements in physical functioning, fatigue, pain, and cognitive performance and provides the rationale for a controlled trial in ME/CFS to confirm therapeutic efficacy. The normalization of thalamic hyperconnectivity following HBOT and its association with clinical response highlights the role of thalamic FC in ME/CFS pathophysiology and underscores the need for larger, controlled trials in ME/CFS to confirm therapeutic efficacy.

## 1. Introduction

Myalgic encephalomyelitis/chronic fatigue syndrome (ME/CFS) is a chronic, debilitating systemic disease involving dysfunction of the neurological, vascular, immune, autonomic, and energy metabolism systems [1], [2], [3], [4], [5], [6], [7]. It is characterized by profound fatigue along with a range of other symptoms including pain, cognitive impairment, autonomic dysfunction, and sleep disturbances [8]. Symptoms are typically worsened by often minor physical or mental exertion referred to as post-exertional malaise (PEM) [8].

ME/CFS affects a significant number of patients, though estimates vary depending on the case definition. The worldwide pre-pandemic prevalence is estimated in the range of 0**·**1% to 0**·**7% [8]. The condition is triggered by an infection in the majority of cases [8]. ME/CFS has been shown in a subset of patients with post-COVID syndrome, and SARS-CoV-2 is now among the most frequent triggers of the disease [9]. Therefore, prevalence is expected to have risen significantly in the aftermath of the COVID-19 pandemic. Despite the high demand for curative options, there is currently no regulatory approved therapeutic option for ME/CFS. Several targeted treatments are currently assessed in clinical trials including antiviral drugs, immunomodulators, and neuromodulators [10]. While some approaches have shown promise in small studies, no treatment has yet demonstrated consistent efficacy across broader patient populations and current guidelines do not recommend the use of these treatments outside of clinical trials [11]. Consequently, there remains a strong need for further research aimed at identifying effective treatments.

Hyperbaric Oxygen Therapy (HBOT) is an established treatment for conditions such as decompression sickness, necrotizing soft tissue infections, carbon monoxide poisoning, and traumatic ischemia, where improving oxygen delivery can aid recovery. HBOT involves breathing 100% oxygen in a pressurized chamber, typically at 1**·**5 to 2**·**5 times normal atmospheric pressure. These conditions permit a significant increment of the oxygen supply to blood and to tissues even without the contribution from hemoglobin [12].

When applying HBOT in multiple sessions, the intermittent exposure to supraphysiological oxygen levels followed by normoxia can trigger many of the same cellular pathways as true hypoxia, an effect summarized as the hyperoxia-hypoxia paradox [13]. Through this effect, HBOT elicits a cascade of cellular responses that collectively may address many pathophysiological aspects of ME/CFS: HBOT may stabilize HIF-1α and thereby lead to an upregulation of enzymes for anaerobic glycolysis to sustain ATP production when oxygen is low, while tempering oxidative stress over time [14]. Simultaneously, HIF-1α can drive the expression of growth factors like vascular endothelial growth factor (VEGF), leading to angiogenesis and improving oxygen delivery [13]. Concurrently, HBOT modulates the immune system by reducing excessive inflammatory cytokines (IL-1, IL-6, TNF-α) and promoting anti-inflammatory profiles (e.g. IL-10, TGF-β) [15], [16].

In ME/CFS patients, in 2013, an uncontrolled trial (n = 16) reported significant reductions in fatigue after 15 HBOT sessions, whereas an earlier pilot study had found no significant improvements in fatigue, pain, or physical functioning [17], [18]. Both trials lacked follow-up assessments, leaving the durability of the effect unknown, and did not evaluate further outcome measures. Following the COVID-19 pandemic however, several studies, including one randomized controlled trial (RCT) and case series, investigated the potential effects of HBOT in post-COVID syndrome, a condition with overlapping symptoms and presumed pathophysiology. Overall, the findings consistently showed improvements in quality of life, fatigue, cognitive function, neuropsychiatric symptoms, and cardiopulmonary function [19]. Most notably, an RCT from 2022 showed improvements in executive functions, psychiatric symptoms, pain and fatigue [20]. These clinical improvements were correlated with increased gray-matter perfusion and microstructural integrity in several brain regions, including the insula and frontal gyrus [21]. A one-year follow-up after the last HBOT session reported lasting effects [22].

Given the overlaps in symptomatology and pathophysiology with post-COVID syndrome, here we aimed to explore the effectiveness and feasibility of HBOT treatment in ME/CFS patients. Effectiveness was evaluated using both pa-tient-reported as well as physician-assessed outcomes. Furthermore, we explored structural and functional brain changes in ME/CFS using MRI to identify potential neural mechanisms associated with therapeutic response. Several resting-state functional MRI (fMRI) studies suggest aberrant connectivity across multiple large-scale neural networks in ME/CFS [23]. In addition, arterial spin labeling (ASL) studies have demonstrated reduced regional cerebral blood flow in patients with ME/CFS, which may be associated with altered functional network dynamics [24], [25]. By integrating clinical and neuroimaging data, this study seeks to clarify the role of HBOT in post-COVID ME/CFS and contribute to the development of evidence-based treatments.

## 2. Materials and Methods

### 2.1 Study design

This prospective cohort study was conducted at the Institute of Medical Immunology, Charité - Universitätsmedizin Berlin, recruited patients between August 2023 and March 2025. Thirty patients completed 40 sessions of HBOT, as well as one baseline and one follow-up visit four weeks after treatment. This study was being conducted within the National Clinical Studies Group (NKSG), a clinical trial and translational research platform focused on developing therapies for post-COVID syndrome and ME/CFS, funded by the German Ministry of Education and Research (BMBF) [10].

### 2.2 Participants

Patients were diagnosed and recruited at the Institute of Medical Immunology at the Charité. The diagnosis of ME/CFS was based on the Canadian Consensus Criteria (CCC) and PEM lasting for a minimum of 14 hours [26], [27]. Patients were excluded from this study if they had relevant comorbidities [28], pre-existing fatigue, evidence of organ dysfunction, or acute or chronic infections such as HIV or hepatitis. Additionally, patients with absolute or relative contraindications for HBOT, including a history of epileptic seizures, pneumothorax, or severe obstructive pulmonary disease, were excluded [12]. Patients who were unable to leave their homes due to the severity of their illness were also excluded.

All patients provided written informed consent prior to study participation. The Ethics Committee of the Charité - Universitätsmedizin Berlin approved this study in accordance with the 1964 Declaration of Helsinki and its later amendments (protocol code EA1/129/23, date of approval: 04 July 2023).

In addition, a group of age- and sex-matched healthy individuals (HCs) was recruited for imaging comparisons. HCs had no history of neurological or psychiatric disorders, chronic fatigue, or any contraindications to MRI. All HCs provided written informed consent.

### 2.3 Procedures

HBOT was administered in a multiplace Starmed-Quadro chamber (HAUX, Germany) at the Center for Hyperbaric Oxygen Therapy and Diving Medicine at the Department of Anaesthesia, Intensive Care Medicine, Emergency Medicine and Pain Therapy of the Vivantes Klinikum im Friedrichshain, Berlin, Germany. Patients received 40 sessions of HBOT over eight to 16 weeks, with up to five sessions per week in an outpatient setting. The HBOT protocol consisted of breathing 100% oxygen via mask at 2 ATA (athmospheres absolute) for 90 minutes, with five-minute air breaks every 20 minutes.

For patient-reported outcomes, questionnaires were completed before HBOT, at four weeks into treatment, on the final day of HBOT, and four weeks after completion, and were validated by physicians. Follow-up assessments continued at bimonthly intervals. Patients’ health-related quality of life was assessed using the 36-Item Short-Form Survey (SF-36), with scores ranging from 0 to 100 and 100 indicating no limitations [29]. Response to HBOT treatment was defined as a minimum increase of 10 points in the SF-36 physical functioning domain (SF-36 PF) from baseline to four weeks post HBOT, indicating a clinically relevant improvement [30]. Fatigue was evaluated using the Chalder Fatigue Scale (CFQ), which ranges from 0 to 33, with a total score of 29 or more suggesting relevant fatigue [31]. Additionally, disease-related disability was scored using the Bell score, which rates restriction in daily functioning on a scale from 0 to 100, with 100 indicating no restriction [32].

Physician-assessed outcomes were recorded at baseline and four weeks after HBOT completion. Handgrip strength of the dominant hand was measured using a digital hand dynamometer (EH101, Deyard, Shenzhen, China). Measurements were conducted with patients sitting upright, facing a standard table. The forearm of the dominant hand was placed on the table in full supination while holding the dynamometer. Under supervision and with verbal encouragement, patients pulled the handle ten times with maximum force for three seconds, followed by a five-second relaxation phase. The dynamometer displayed the highest value reached within each repetition measured in kg. The highest recorded value across ten repetitions was noted as the maximum strength (Fmax), and the average strength (Fmean) of each session was calculated [33].

Orthostatic dysfunction was assessed using a passive standing test. During the test, the patient’s blood pressure, heart rate, and any reported symptoms were recorded at one-minute intervals. Resting measurements were taken in a supine position. Subsequently, patients were asked to stand straight with their shoulders leaned against a wall and their heels one step away from the wall for ten minutes. The test was terminated early if the patient was unable to continue standing due to severe orthostatic dysfunction [34].

The symbol digit modalities test (SDMT) was used to assess information processing speed and efficiency. Patients were timed to determine the number of correct responses they could complete within 90 seconds. The absolute number of correct responses was recorded, along with standard deviations adjusted for age, gender, and education status, as specified in the test manual [35].

The 1-minute sit-to-stand test was used to assess exercise capacity. The test was conducted using a standard chair. Patients were asked to sit forward with their feet flat on the floor and arms crossed over the chest. The total number of complete sit-to-stand cycles within one minute was recorded as the final score. While resting during the test was allowed, the test was terminated early if the patient was unable to continue, and the total number of completed sit-to-stand cycles up to that point was then recorded as the final score [36].

Study data were collected and managed using the REDCap electronic data capture tools hosted at Charité - Universitätsmedizin Berlin [37].

### 2.4 MRI data acquisition

MRI data were acquired at the Berlin Center of Advanced Neuroimaging (BCAN) using a Siemens 3T PRISMA scanner with a 64-channel head coil (Siemens, Erlangen, Germany). A high-resolution three-dimensional T1-weighted MRI sequence (3D-MPRAGE; TR = 2500ms, TE = 2·64ms; TI = 1000 mm, voxel size 1·0 mm^3^) and a ten-minute resting-state functional MRI (fMRI) scan (720 volumes, TR = 0·8sec, TE = 37 ms, voxel size = 2·0 mm^3^) were acquired. Patients underwent a baseline MRI session within four weeks prior to HBOT therapy and a follow-up session within four weeks after the conclusion of therapy. HCs underwent a single MRI scan. Both patients and HCs followed the same scanning protocol.

### 2.5 MRI data analysis

Whole-brain volumetric analyses were performed using the FreeSurfer image analysis suite (version 7.4.1) [38]. Resting-state fMRI data preprocessing was conducted using fMRIPrep (version 23.2.0) [39], including skull-stripping, co-registration, normalization, unwarping, noise component extraction, and segmentation. In addition, functional data were smoothed (6 mm Gaussian kernel) and denoised in the CONN Toolbox (version 22, update v24.07) [40]. Denoising included regression of confounding effects from white matter and cerebrospinal fluid components (10 CompCor components each [41]), motion parameters and their first-order derivatives (12 regressors), and outlier volumes. The denoised BOLD signal was then band-pass filtered between 0·008 and 0·09 Hz.

Seed-based functional connectivity (FC) analyses were also performed in CONN. Regions of interest (ROIs) included the bilateral thalamus (Harvard-Oxford subcortical structural atlas) and thalamic subregions [42] (connectivity-based parcellation by Boeken et al [43]. Original masks were transformed to each subject’s functional space and used without modification as seed ROIs. Three main comparisons were conducted: (1) patients pre-treatment vs. HCs, (2) patients post-treatment vs. HC, and (3) patients post-treatment vs. pre-treatment. Age and sex were included as covariates in the comparisons with HC but not in the within-patient comparison. Additionally, correlation analyses were conducted to examine associations between pre- and post-treatment functional connectivity changes and clinical outcome measures. These measures included the main outcome “response to HBOT treatment” as well as fatigue severity (assessed with the CFQ), disease-related disability, handgrip strength, SDMT scores, and exercise capacity (assessed using the 1-minute sit-to-stand test).

### 2.6 Statistical analysis

Statistical analyses were conducted using R version 4.3.0 and RStudio version 2023.03.1. A linear mixed-effects model (LMM) was employed to assess changes in patient-reported variables across different time points. This analysis was performed using the lmer function from the lme4 package (version 1.1-35.5), and ggplot2 (version 3.5.0) was used for data visualization. For each outcome variable, the LMM included time as a fixed effect and patient number as a random effect to account for within-patient correlation. The mixed model was fitted using restricted maximum likelihood (REML), and statistical significance was evaluated using t-tests with p-values approximated through Satterthwaite’s method for degrees of freedom, implemented via the lmerTest package (version 3.1-3). Missing data were accounted for by using all available observations in the model, allowing for the estimation of fixed and random effects without listwise deletion, assuming data were missing at random.

For group comparisons, the non-parametric Mann-Whitney U test was used for unpaired data, and the Wilcoxon signed-rank test was used for paired data. Group comparisons for categorical data were performed using Fisher’s exact test and corrected for multiple comparisons using the Bonferroni method. Correlation analysis was performed using the nonparametric Spearman coefficient. Effect sizes were reported throughout as standardized mean differences (Hedges’ g) with a g of 0·2 is considered a small, 0·5 a medium and 0·8 a large effect. A two-tailed p-value of <0·05 was considered to indicate statistical significance.

To identify baseline predictors of treatment response, exploratory logistic regression analyses were performed with responder status as the dependent variable. Baseline SF-36 general health, CFQ physical fatigue, and SDMT performance were entered as independent variables.

For imaging data, second-level general linear model (GLM) analyses were conducted in the CONN Toolbox to assess group differences and within-subject changes in FC. Statistical significance for group comparisons of imaging results was determined using a voxel-wise threshold of p < 0.001 and a cluster-level threshold of p < 0·05, both corrected for multiple comparisons to control the false discovery rate (FDR). For within-subject comparisons, a less stringent voxel-wise threshold of p < 0·01 was also applied, with the same cluster-level FDR correction, to account for the smaller sample size.

## 3. Results

### 3.1 Patient characteristics

A total of 253 patients who met the CCC for post-infectious ME/CFS and had consented to be contacted for clinical trials were screened for study participation between August 2023 and March 2025. Of these, 39 patients living more than 100 km from the study site were excluded for practical reasons. Among the remaining 214 patients, 95 responded and expressed interest in participating. Of these 95 patients, 37 were included in this study. Another 30 were offered to participate in a follow-up study with shorter treatment of 20 HBOT sessions and 28 were excluded due to contraindications for HBOT or because they self-assessed their illness as too severe to attend regular appointments outside their home.

Of the remaining 37 patients, 30 underwent 40 sessions of HBOT and comprise the treatment cohort analyzed in this study (Fig. 1). Seven patients were male, and 23 were female, with a mean age of 42·33 ± 11·73. In 27 patients (90%), SARS-CoV-2 infection was identified as the disease trigger, while the remaining patients had other viral triggers. The mean disease duration at study inclusion was 27·03 ± 11·21 months. All patients had a severe to moderate degree of disability, with Bell scores ranging from 30 to 70. Additional patient characteristics are presented in Table 1. Thirteen patients had comorbid POTS, and eight had a preexisting autoimmune condition.

**Figure 1:**
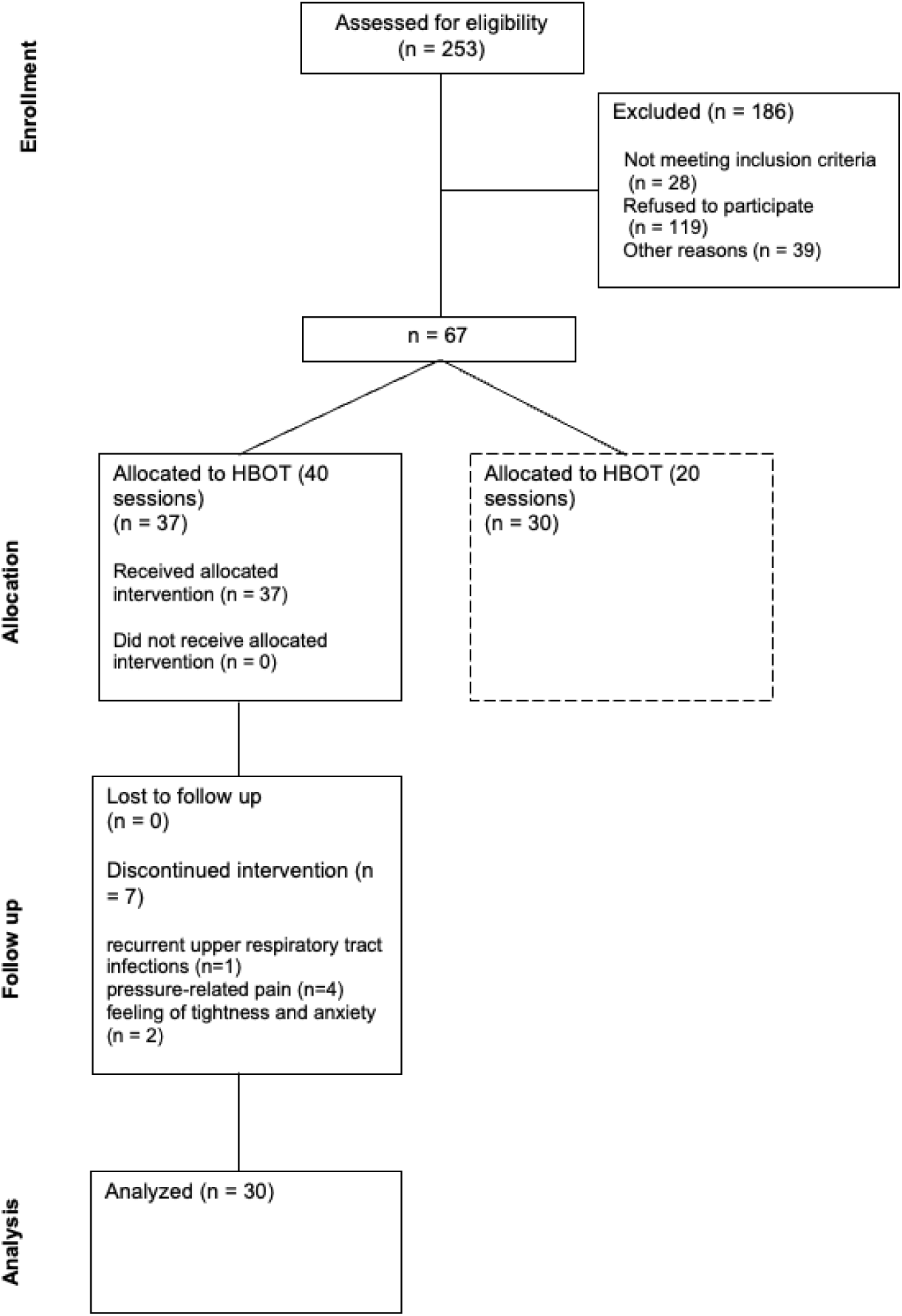
Consort diagram of patients eligible, recruited, followed up, and included in analysis.

**Table 1:** Patient characteristics at study inclusion

| Characteristics |  |
| --- | --- |
| **Total patients (n)** | 30 |
| Male (n, %) | 7 (23.33%) |
| Female (n, %) | 23 (76.67%) |
| **Age, years (mean ± SD)** | 42.33 ± 11.73 |
| **BMI, kg/m<sup>2</sup> (mean ± SD)** | 24.45 ± 4.04 |
| **Education level (n, %)** |  |
| No degree | 0 (0%) |
| High School | 4 (13.33%) |
| Vocational Training | 10 (33.33%) |
| University degree | 16 (53.33%) |
| **Disease duration, months (mean ± SD)** | 27.03 ± 11.21 |
| **Infectious trigger (n, %)** | 30 (100%) |
| COVID-19 | 27 (90%) |
| EBV | 1 (3.33%) |
| Upper respiratory tract infection | 2 (6.67%) |
| **Diagnostic criteria used (n, %)** |  |
| CCC | 30 (100%) |
| IOM | 30 (100%) |
| **SF-36 PF score (mean ± SD)** | 37.50 ± 20.63 |
| **Bell Score (mean ± SD)** | 38.00 ± 10.95 |
| **Handgrip strength, kg (mean ± SD)** |  |
| Mean handgrip strength (Fmean) | 15.82 ± 11.03 |
| Maximum handgrip strength (Fmax) | 19.02 ± 12.02 |
| **Comorbidities (n, %)** |  |
| POTS | 13 (43.33%) |
| Immunodeficiency | 8 (26.67%) |
| CD4 lymphocytopenia | 1 (3.33%) |
| IgM deficiency | 2 (6.67%) |
| IgA deficiency | 1 (3.33%) |
| MBL deficiency | 5 (16.67%) |
| Autoimmune condition | 8 (26.67%) |
| Hashimoto's thyroiditis | 6 (20%) |
| Alopecia areata | 1 (3.33%) |
| Graves' disease | 1 (3.33%) |
| Celiac disease | 2 (6.67%) |
| **Lifestyle Factors** |  |
| Smoking/nicotine use (n, %) | 1 (3.33%) |
| Alcohol use (n, %) | 10 (33.33%) |
| **Previous Treatments (n, %)** |  |
| Inpatient rehab | 15 (50%) |
| Low dose naltrexone (LDN) | 8 (26.67%) |
| Ivabradine | 6 (20%) |
| Pyridostigmine | 4 (13.33%) |
| Antidepressants | 9 (30%) |
| Antihistamines | 9 (30%) |

BMI: Body mass index, CCC: Canadian Consensus Criteria; COVID-19: Coronavirus disease 2019; EBV: Epstein-Barr virus, IOM: Institute of Medicine, POTS: Postural tachycardia syndrome, SF-36 PF: Short Form-36 Health Survey - Physical Functioning.

### 3.2 Treatment course, adverse events, and response

A total of 37 patients were enrolled. One patient was unable to complete the treatment due to recurrent upper respiratory tract infections, while six patients had to discontinue early due to pressure-related pain in the sinuses and ears (n = 4) or a feeling of tightness and anxiety (n = 2). Accordingly, 30 patients completed 40-sessions of HBOT within eight to 16 weeks, the average time to complete all 40 sessions was 13 weeks. Patients often required breaks between sessions to prevent symptom exacerbation.

After completing the 40 HBOT sessions, 23 patients (76·67%) reported subjective improvement, and 26 patients (86.67%) reported that they would like to receive HBOT again in the future. The most commonly reported side effects included a feeling of pressure or mild ear pain (n = 13, 43·33 %) and reversible myopia (n = 17, 56·67%). None of the patients reported severe or permanent side effects, and none had to discontinue the treatment due to PEM or worsening of other ME/CFS symptoms.

As shown in Figure 2a, the greatest improvement in SF-36 PF was observed on the last day of HBOT, with a mean increase of 6·3 points (CI: 1·98–10·68, p = 0·006) in the cohort of 30 patients. This corresponds to a standardized mean difference of Hedges’ g = 0·71 (95% CI: 0·21–1·21), indicating a moderate to large effect. Four weeks after HBOT completion, a significant mean increase of 4.5 points compared to baseline (CI: 0·15–8·85, p = 0·047) was documented, corresponding to a standardized mean difference of Hedges’ g = 0·51 (95% CI: 0·01–1·01) indicating a moderate effect. A clinically meaningful improvement defined as an increase of at least 10 points in SF-36 PF [29], was observed in 11 out of 30 patients (36·67%) 4 weeks post-HBOT, with the highest individual improvement reaching 35 points.

**Figure 2:**
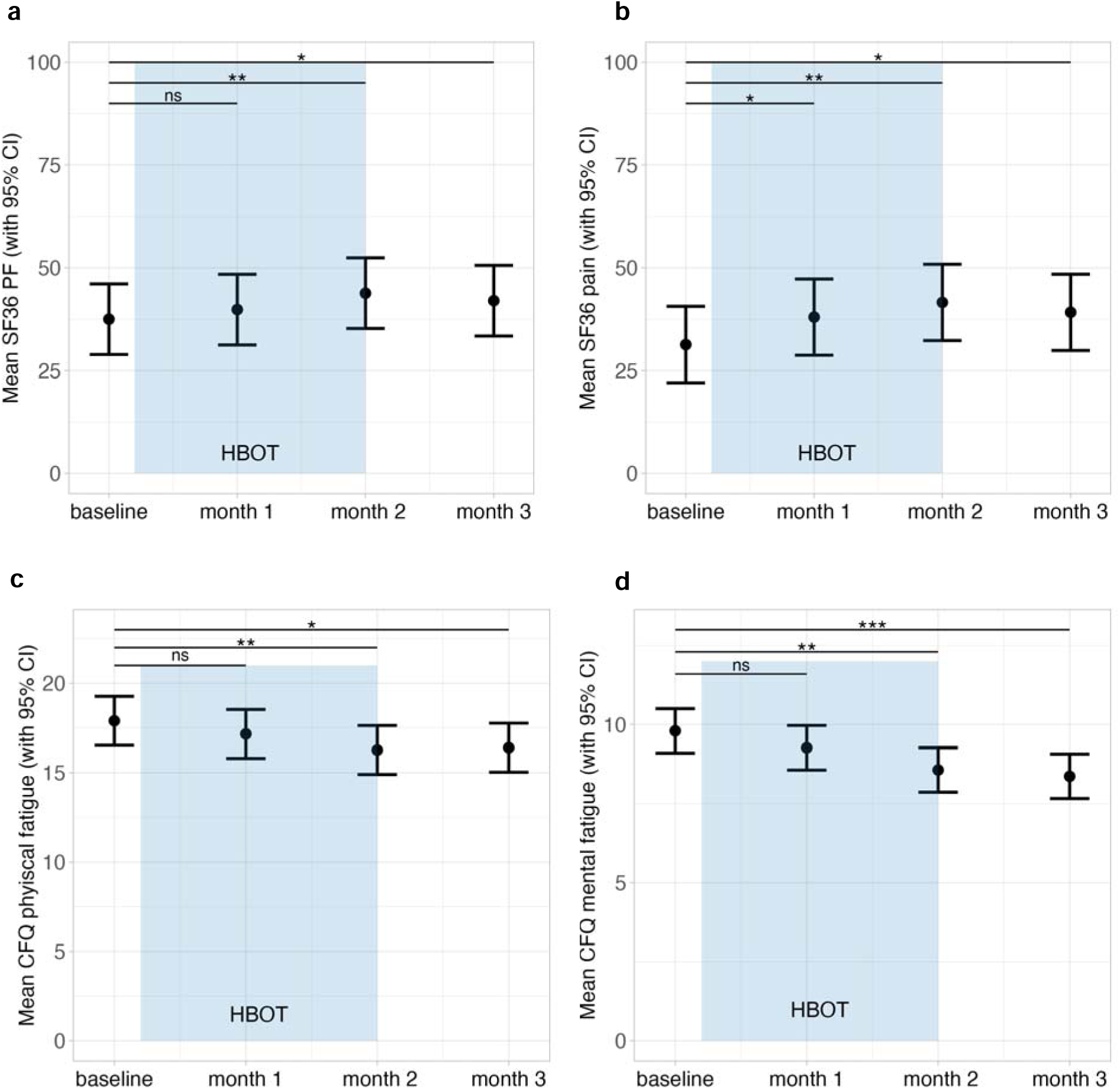
Course of patient-reported symptoms over the study period. The duration of HBOT therapy is indicated by the blue bar. Error bars represent 95% confidence intervals. Data were analyzed using a linear mixed-effects model fitted by restricted maximum likelihood (REML), with t-tests computed using Satterthwaite’s method for degrees of freedom. Significance levels are indicated as ^∗^p < 0·05, ^∗∗^p < 0·01, ^∗∗∗^p < 0·001. The panels display the trajectories of: a) Mean 36-Item Short-Form Survey physical functioning domain (SF-36 PF) scores over the study period. A higher score indicates less restriction in physical functioning. b) Mean 36-Item Short-Form Survey pain domain (SF-36 pain) scores over the study period. A higher score indicates less pain. c) Mean Chalder Fatigue Scale physical fatigue domain (CFQ physical fatigue) scores over the study period. A higher score indicates more severe fatigue. d) Mean Chalder Fatigue Scale mental fatigue domain (CFQ mental fatigue) scores over the study period. A higher score indicates more severe fatigue.

Patients reported improvements in several core clinical symptoms following HBOT, including a significant reduction in fatigue and pain. The total CFQ fatigue score decreased, with the greatest improvement observed 4 weeks post-HBOT (-2·93 points; CI: -4·54 to -1·33, p < 0·001), corresponding to a standardized mean difference of Hedges’ g = - 0·89 (95% CI: -1·39 to -0·39) indicating a strong effect. Individual scores for mental and physical fatigue both significantly improved, as presented in Figure 2c and Figure 2d. Pain, measured by the SF-36 pain domain, improved with a maximum increase of 10·25 points (CI, 3·86–16·64, p = 0·002) occurring on the last day of HBOT, corresponding to a standardized mean difference of Hedges’ g = 0·79 (95% CI: 0·29–1·28). These improvements remained significant through month 1, as shown in Figure 2b.

### 3.3 Functional tests

To further quantify and objectively assess the self-reported improvement, we conducted several functional tests during the study visits at baseline and 4 weeks post-HBOT. Processing speed, as measured by the SDMT, significantly improved after HBOT, increasing from a median of -1·07 SD (IQR: -1·7 to -0·25; SD; values >-1.0 indicating reduced processing speed) to a median of -0·7 SD (IQR: -1·4 to 0·2 SD) (V = 91, p = 0·011) after HBOT, corresponding to a standardized mean difference of Hedges’ g = 0·52 (95% CI: 0·15–0·89) reflecting a medium size effect (Figure 3a). The improvement in SDMT scores was comparable between responders and non-responders on the SF-36 PF subscale (Table 2).

**Figure 3:**
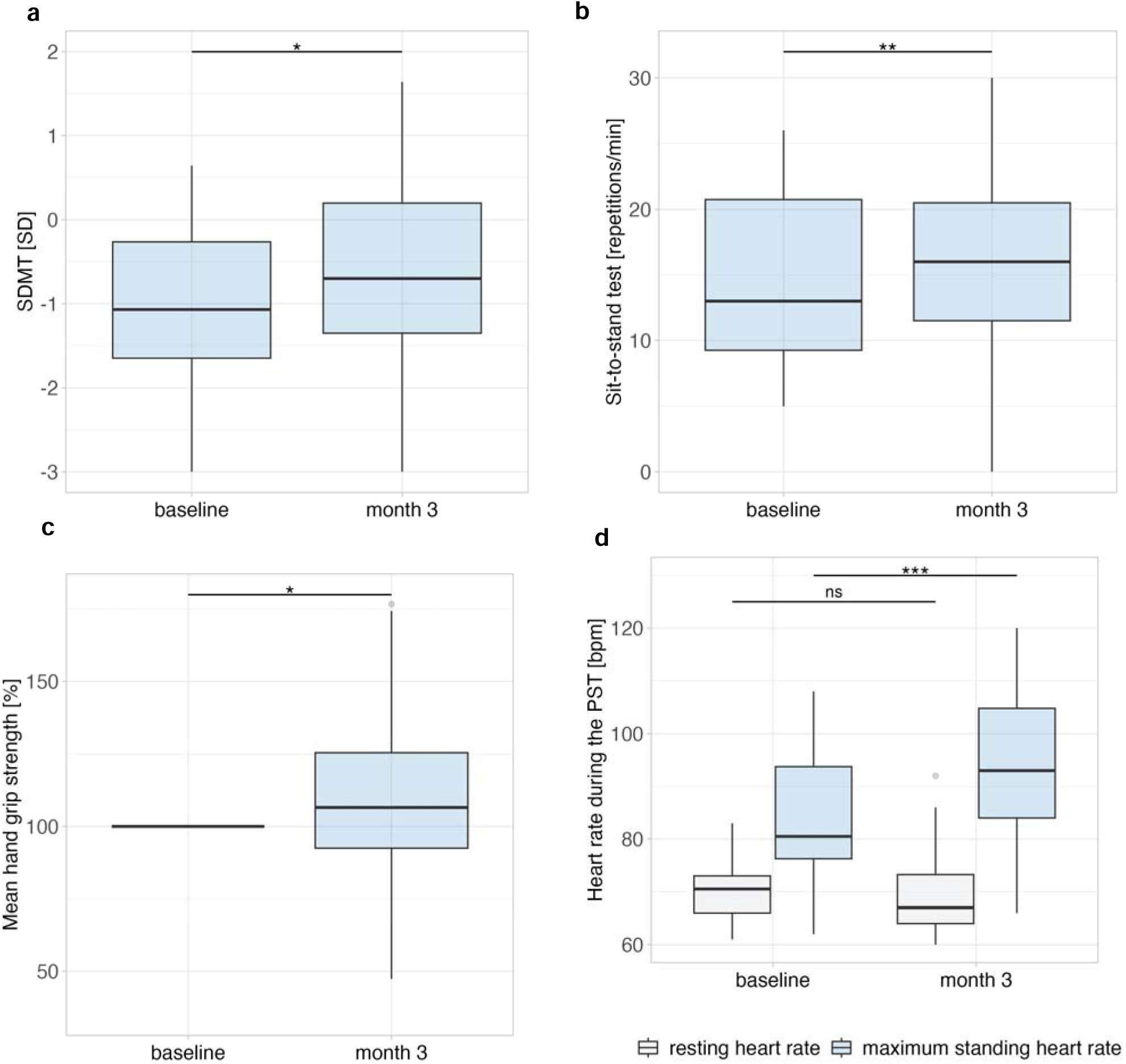
Physician-assessed parameters at baseline and one month post-HBOT. Data were analyzed using the paired Wilcoxon signed-rank test, with significance levels indicated as ^∗^*p* < 0·05, ^∗∗^*p* < 0·01, ^∗∗∗^*p* < 0·001. The panels display the results of: a) Symbol Digit Modalities Test (SDMT) reported as standard deviations adjusted for age, gender, and education status, as specified in the test manual [35]; b) repetitions in the 1 minute sit-to-stand test; c) mean handgrip strength over ten repetitions (Fmean), shown as the percentage increase; d) resting supine heart rate and maximum standing heart rate in beats per minute (bpm) during the passive standig test.

**Table 2:** Patient-reported outcomes and functional test results before and 4 weeks after HBOT in responders (n = 11) and non-responders (n = 19). Responders were defined as patients with an improvement of ≥10 points in SF-36 physical functioning. Patient-reported outcomes were analyzed using a linear mixed-effects model fitted by restricted maximum likelihood (REML), with t-tests computed using Satterthwaite’s method for degrees of freedom with significance levels indicated as ^∗^*p* < 0·05, ^∗∗^*p* < 0·01, ^∗∗∗^*p*< 0·001. Functional tests were analyzed using the paired Wilcoxon Signed Rank Test with significance levels indicated as ∗p < 0.05, ∗∗p < 0.01, ∗∗∗p < 0.001.s

|  | Responders (n=11) |  | non-responders (n=19) |  |
| --- | --- | --- | --- | --- |
| Characteristics | baseline | month 3 | baseline | month 3 |
| **SF-36 score (mean ± SD)** |  |  |  |  |
| Physical functioning | 33.18 ± 21.48 | 50.91 ± 26.25 *** | 40.00 ± 20.28 | 36.84 ± 22.68 |
| Role limitations due to physical health | 4.55 ± 10.11 | 18.18 ± 33.71 | 1.32 ± 5.74 | 1.32 ± 5.74 |
| Role limitations due to emotional health | 75.76 ± 42.40 | 96.97 ± 10.05 | 54.39 ± 43.33 | 33.33 ± 47.14 * |
| Energy/fatigue | 20.00 ± 20.86 | 28.64 ± 18.72 | 13.42 ± 15.28 | 14.74 ± 15.50 |
| Emotional well-being | 62.55 ± 21.41 | 69.45 ± 17.28 | 53.26 ± 21.99 | 54.74 ± 20.91 |
| Social functioning | 27.27 ± 20.78 | 48.86 ± 33.75 ** | 23.68 ± 18.11 | 17.11 ± 20.07 * |
| Pain | 37.50 ± 21.99 | 47.73 ± 24.12 | 27.76 ± 20.27 | 34.21 ± 28.92 |
| General health | 28.18 ± 14.37 | 32.73 ± 19.45 | 24.74 ± 13.49 | 24.21 ± 14.65 |
| **CFQ score(mean ± SD)** | 26.73 ± 5.08 | 21.45 ± 6.09 ** | 28.26 ± 3.16 | 26.68 ± 4.33 |
| Physical fatigue | 16.64 ± 3.61 | 13.73 ± 4.47 * | 18.63 ± 2.43 | 17.95 ± 3.46 |
| Mental fatigue | 10.09 ± 1.70 | 7.73 ± 2.69 ** | 9.63 ± 1.46 | 8.74 ± 1.52 |
| **Bell score (mean ± SD)** | 39.09 ± 13.00 | 40.91 ± 17.58 | 37.37 ± 9.91 | 36.32 ± 12.12 |
| **Handgrip strength, kg, % (mean ± SD)** |  |  |  |  |
| Mean handgrip strength (Fmean) | 17.28 ± 9.56<br>100% | 18.98 ± 10.18<br>115.10% ± 27.31% | 14.97 ± 11.96<br>100% | 15.46 ± 11.56<br>117.15% ± 46.06% |
| Maximum handgrip strength (Fmax) | 21.14 ± 11.29<br>100 % | 21.67 ± 10.97<br>111.26% ± 30.41% | 17.80 ± 12.56<br>100 % | 19.04 ± 12.52<br>117.37% ± 38.05% |
| **SDMT, SD (mean ± SD)** | -1.01 ± 1.03 | -0.79 ± 1.13 | -1.05 ± 1.10 | -0.61 ± 1.35 * |
| **1-minute sit-to-stand, repetitions (mean ± SD)** | 14.40 ± 8.26 | 21.45 ± 14.05 ** | 16.35 ± 8.60 | 17.37 ± 10.58 |

| Passive standing test heart rate, bpm (mean ± SD) | | | | |
| --- | --- | --- | --- | --- |
| Maximum standing heart Rate | 82.82 ± 12.01 | 92.00 ± 17.01 * | 84.63 ± 12.02 | 93.26 ± 13.65 * |
| $\Delta$ Heart rate (supine to standing) | 18.27 ± 8.79 | 26.82 ± 11.59 * | 19.22 ± 11.10 | 23.89 ± 9.93 * |

CFQ: Chalder Fatigue Questionnaire, SDMT: symbol digit modalities test, SF-36: Short Form-36 Health Survey

Performance in the 1-minute sit-to-stand test significantly improved from the baseline visit (median = 14 repetitions, IQR: 9–22) to the follow-up visit (median = 17·5 repetitions, IQR: 12–22) (V = 52, p = 0·001) (Figure 3b). This corresponds to a standardized mean difference of Hedges’ g = 0·66 (95% CI: 0·25–1·07), indicating a moderate effect. Mean handgrip strength (Fmean) significantly improved from baseline to the visit 4 weeks post-HBOT, with a median increase of 1·14 kg. When calculating the percentage increase for each patient individually, the median improvement in Fmean was 106% (IQR: 92·49–131·7%) (V = 136, p = 0·048), equating to Hedges’ g = 0·40 (95% CI: 0·04–0·76) (Figure 3c), and 106% (IQR: 98·97–135·3%) for the maximum handgrip strength (Fmax) (V = 117, p = 0·031). This corresponded to a standardized mean difference of Hedges’ g = 0·42 (95% CI: 0·05–0·78), indicating a small-to-moderate effect.

While there was no significant change in resting supine heart rate, we observed a significant increase in the maximum standing heart rate after HBOT (V = 58, p < 0·001, Hedges’ g = 0·65 (95% CI: 0·26–1·03)). The median supine to standing heart rate difference increased from 16 bpm (IQR: 11–25) baseline to 24 bpm (IQR: 17–31) at 4 weeks post-HBOT (V = 59, p = 0·002, Hedges’ g = 0·65 (95% CI: 0·25–1·04) ). Resting and maximum heart rates during the passive standing test are shown in Figure 3d.

### 3.4 Predictive parameters for response

There were no significant differences in any of the patient characteristics presented in Table 1 between patients who responded to the treatment (defined as an increase of at least 10 points in SF-36 PF) and those who did not. Results for patient-reported outcomes as well as functional test results before and 4 weeks after HBOT in responders (n = 11) and non-responders (n = 19) are summarized in Table 2.

Greater improvement in SF-36 PF at 4 weeks post-HBOT was significantly associated with better baseline SF-36 general health (r = 0·38, p = 0·039), lower baseline CFQ physical fatigue (r = -0·39, p = 0·033), and better baseline SDMT performance (r = 0·36, p = 0·048). To explore predictors of treatment response in SF-36 PF, logistic regression analyses were conducted with responder status (n = 11 responders, n = 19 non-responders) as the dependent variable. In univariate analyses, lower baseline CFQ physical fatigue scores showed a trend toward predicting response (OR = 0·79, 95% CI: 0·58–1·02; p = 0.096), whereas baseline SF-36 general health (OR = 1·02, 95% CI: 0·96–1·08; p = 0.50) and baseline SDMT performance (OR = 1·04, 95% CI: 0·51–2·18; p = 0·92) were not associated with response. In the multivariable model including all three predictors, lower baseline CFQ physical fatigue remained the strongest predictor, although the association did not reach statistical significance (OR = 0·76, 95% CI: 0·50–1·03; p = 0·11). The overall discriminative ability of the model was modest (AUC = 0.66).

### 3.5 Brain imaging analyses

Volumetric analysis revealed no significant differences in whole-brain or regional volumes between patients pre-treatment and HCs, between patients post-treatment and HCs, or between patients pre- and post-treatment.

Baseline seed-to-voxel FC analysis showed increased thalamic connectivity with bilateral motor and somatosensory regions as well as bilateral visuo-occipital regions in ME/CFS patients compared to HCs. Specifically, clusters of increased thalamic connectivity were observed with the bilateral precentral and postcentral gyri and the left superior parietal lobule (p < 0·001, t = 5·48, FDR-corrected), the right lateral occipital cortex and lingual gyrus (p < 0·001, t = 5·4, FDR-corrected), and the left occipital pole, lateral occipital cortex, and cuneal cortex (p < 0·001, t = 4·86, FDR-corrected) (see Figure 4a). At follow-up, no significant differences in FC between patients and HCs were observed.

**Figure 4:**
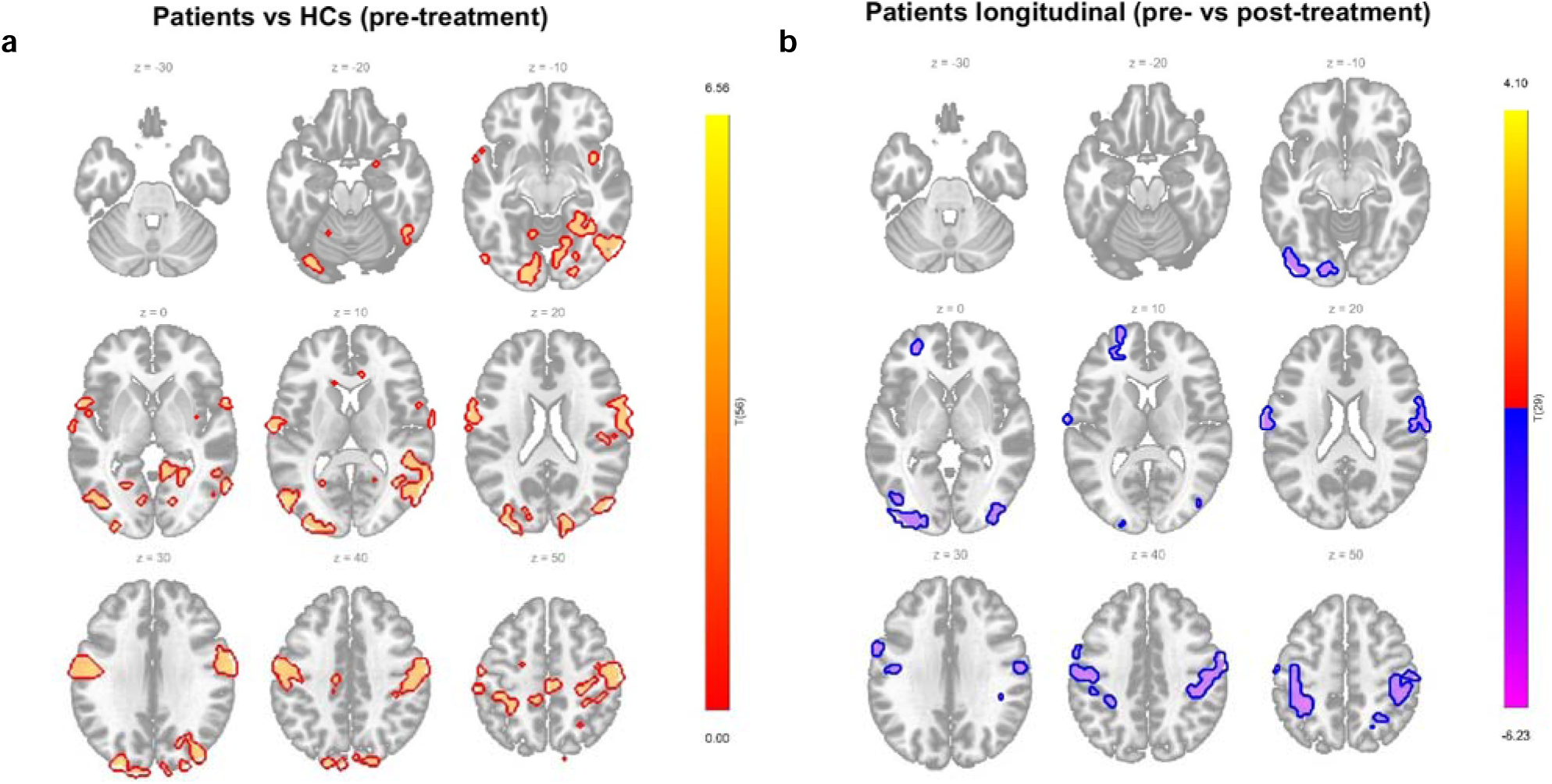
Thalamic functional connectivity in ME/CFS patients Seedlllto&#x2370;voxel functional connectivity maps are displayed as t&#x2370;maps on the MNI&#x2370;152 template. Warm colors indicate stronger positive connectivity; cool colors indicate reduced connectivity. **(a)** Patients vs HCs pre-treatment, seed: bilateral thalamus. Patients showed significantly increased thalamic connectivity with bilateral sensorimotor regions (precentral and postcentral gyri, superior parietal lobule) and bilateral visuo-occipital regions (lateral occipital cortex, lingual gyrus, cuneus, occipital pole). No significant differences were observed post-treatment compared to HCs. Threshold: voxel-wise p < 0·001; cluster-level p < 0·05, FDR-corrected. **(b)** Patients longitudinal (prevs post-treatment), seed: left thalamic sensorimotor cluster. Post-treatment reductions in thalamic connectivity were observed with bilateral sensorimotor regions (precentral and postcentral gyri), left superior parietal lobule, bilateral supramarginal gyri, and left visuo-occipital regions (lateral occipital cortex, occipital pole, occipital fusiform gyrus). Threshold: voxel-wise p < 0·01; cluster-level p < 0·05, FDR-corrected.

Given the functional segregation of the thalamus, an additional analysis was conducted to investigate thalamic subregions. Longitudinal analysis revealed a significant reduction in FC from pre- to post-treatment in patients, specifically between the left sensorimotor cluster of the thalamus and bilateral sensorimotor regions (precentral gyri and postcentral gyri, supramarginal gyrus and superior parietal lobule), as well as the left visuo-occipital regions (lateral occipital cortex, occipital pole, and occipital fusiform gyrus) (right sensorimotor/parietal cluster: t = −6·05; left sensorimotor/parietal: t = −6·70; left occipital: t = −5·56; all p < 0·001, FDR-corrected) (Figure 4b).

### 3.6 Correlation between connectivity changes and clinical outcomes

Subsequently, we examined thalamic FC in physical function responders (n = 11; defined by a ≥10-point increase in SF-36 PF score) compared to non-responders (n = 19).

In responders, connectivity was significantly reduced from pre- to post-treatment between the left thalamic sensorimotor cluster and the left superior parietal lobule, lateral occipital cortex, supramarginal gyrus, postcentral gyrus, and occipital pole (t = −7·54), as well as with the right lateral occipital cortex, superior parietal lobule, and postcentral gyrus (t = −5·21). Reduced connectivity was also observed between the left thalamic sensorimotor cluster and the left occipital pole, lingual gyrus, and occipital fusiform gyrus (t = −4·79) (all p < 0·01, FDR-corrected; Figure 5a).

**Figure 5:**
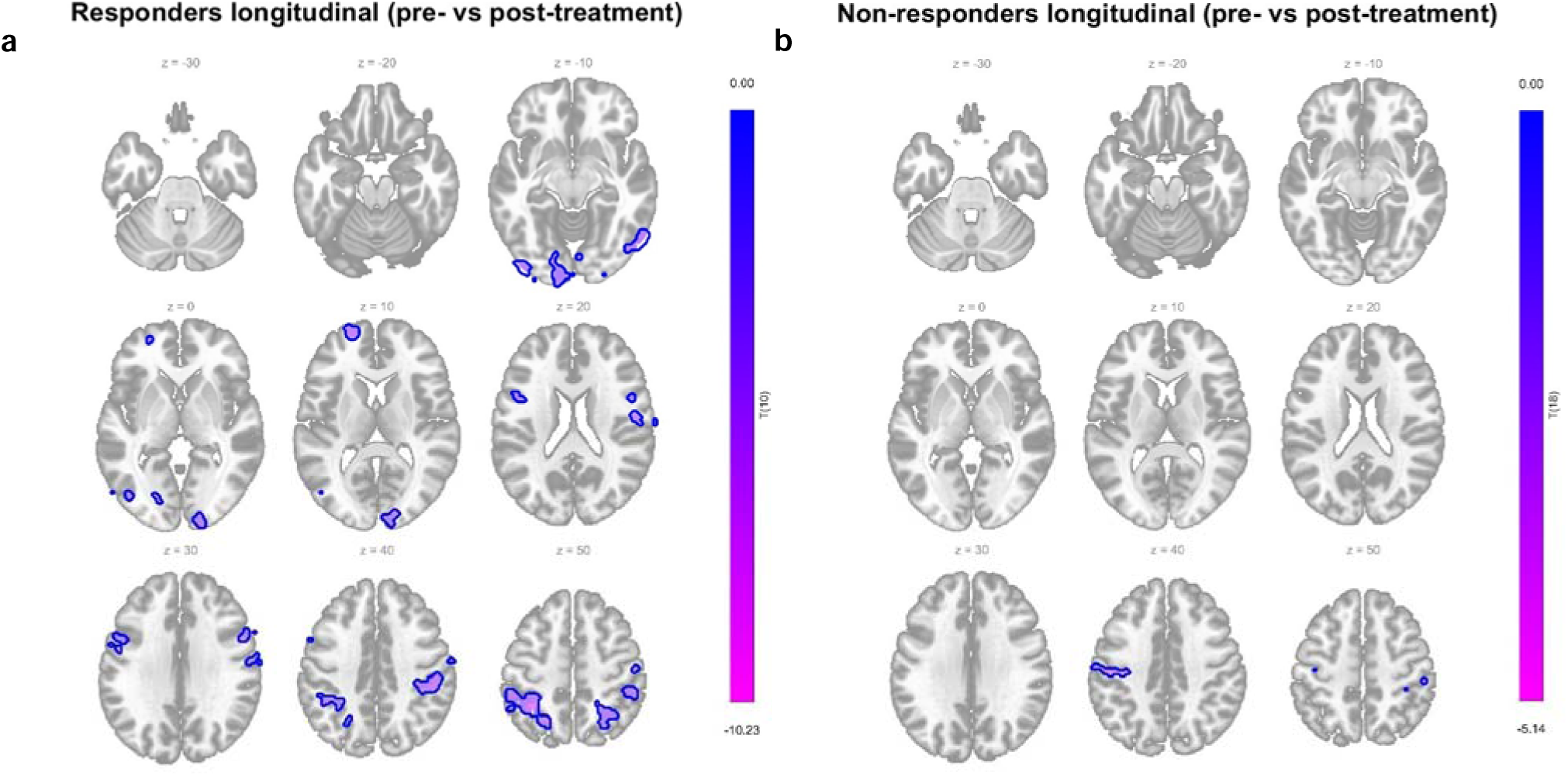
Functional connectivity differences between clinical responders and non-responders Seed-to-voxel functional connectivity maps are displayed as t-maps over the MNI-152 template. Cool colors indicate reduced connectivity. **(a)** Responders, longitudinal analysis (pre-vs post-treatment), seed: left thalamic sensorimotor cluster. Connectivity was reduced from pre- to post-treatment in the left superior parietal lobule, lateral occipital cortex, supramarginal gyrus, postcentral gyrus, and occipital pole, as well as the right lateral occipital cortex, superior parietal lobule, and postcentral gyrus. **(b)** Non-responders, longitudinal analysis (pre- vs post-treatment), seed: left thalamic sensorimotor cluster. Connectivity was reduced from pre- to post-treatment in the bilateral precentral and postcentral gyri. Threshold: voxel-wise p < 0·01; cluster-level p < 0·05, FDR-corrected.

In non-responders, significant FC reductions were limited to small clusters in the right precentral and postcentral gyri (t = −6·44) and the left precentral and postcentral gyri (t = −5·55) (all p < 0.01, FDR-corrected; Figure 5b).

No significant group differences were observed at baseline. At follow-up, responders exhibited greater reductions in connectivity relative to non-responders between the left sensorimotor cluster and both visuo-occipital and sensorimotor regions. These included the right occipital pole and lateral occipital cortex (t = −4·34), the right superior parietal lobule and postcentral gyrus (t = −5·18), and the left lateral occipital cortex, superior parietal lobule, and postcentral gyrus (t = −4·71) (all p < 0·01, FDR-corrected). No significant associations were found between changes in thalamic connectivity and other clinical variables.

## 4. Discussion

This observational study suggests that HBOT may improve physical functioning, reduce pain and fatigue severity, and enhance exercise capacity, muscle strength, and information processing speed in a subset of ME/CFS patients.

To date, only two small studies, in 2003 and 2013, have specifically evaluated HBOT in ME/CFS patients, yielding conflicting results [17], [18]. Recently, interest in HBOT has re-emerged as a potential treatment for post-COVID syndrome [19]. Most notably, a sham-controlled RCT in post-COVID patients demonstrated significant improvements in cognitive function and fatigue, accompanied by parallel improvements in brain perfusion and microstructure [20], [21]. While these results are broadly consistent with our findings, notable differences exist between study populations and treatment protocols. Both protocols employed 40 sessions of 90 minutes each at 2 ATA [20], however, the RCT included five sessions per week and enrolled patients of whom only 77% reported fatigue. ME/CFS patients are generally more severly ill and suffer considerably from post-exertional malaise with the risk of worsening of symptoms due to traveling to the treatment center or sitting upright during therapy. To accommodate this, we implemented a flexible therapy duration and reduced session frequency, ranging from eight to 16 weeks rather than a fixed eight-week schedule. This adjustment resulted in high therapy adherence and no early terminations due to PEM. A small number of patients discontinued treatment early due to pressure-related issues in the sinuses and middle ear. Given that the vast majority of participants reported subjective improvements and expressed interest in receiving HBOT again, we consider this treatment both feasible and acceptable for ME/CFS patients.

Improvement in SF-36 PF at four weeks post-HBOT was the primary outcome of our study. Self-reported physical functioning improved both during treatment and at follow-up, with effect sizes in the moderate-to-large range. 11 out of the 30 (37%) patients experienced a clinically meaningful physical function improvement, defined as an increase of at least 10 points. To complement patient-reported outcomes, we conducted physician-assessed measurements of physical performance, including the 1-minute sit-to-stand test and handgrip strength measurements, both showing significant improvements from pre- to post-HBOT. Improvement in the 1-minute sit-to-stand test was observed only in the group that also reported a clinical meaningful improvement in the SF36 PF, confirming this finding. While the aforementioned ME/CFS trials did not include direct assessments of exercise capacity or muscle strength, findings from post-COVID studies suggest improved exercise capacity post-HBOT, including anecdotal reports of increased 6-minute walk distance and significant improvements in 2-minute step and 30-second sit-to-stand tests [19].

In our study, responders in SF36PF also demonstrated a significant reduction in fatigue[31]. Comparable results were reported in a case series of ten post-COVID patients treated with only ten HBOT sessions, with a large effect size on the CFQ [44]. The same study also reported significant improvements in information processing speed and verbal memory, cognitive domains comparable to those assessed by the SDMT, which likewise improved significantly in our cohort. Interestingly, in our study the improvement in SDMT scores was comparable between responders and non-responders on the SF-36 PF subscale, suggesting that cognitive gains may occur independently of improvements in physical function and fatigue. Similary, pain, as measured by the SF-36 pain domain, improved after HBOT with comparable changes in both groups. Consistent with our findings, post-COVID HBOT studies have reported significant pain relief [20], [45].

In summary, we observed therapeutic effects on several core symptoms in ME/CFS patients, closely resembling those reported in post-COVID patients, despite differences in the treatment duration and frequency of the HBOT protocols used across studies. Findings from our study also suggest heterogenous response patterns, with some patients improving in physical function, fatigue, cognition and pain, while in others improvement was limited to cognition and pain.

Notably, in the passive standing test, patients showed an increased maximum heart rate upon standing, while resting heart rate remained unchanged. This may suggest that HBOT induces subtle changes in cardiovascular regulation improving baroreflex sensitivity and the autonomic responsiveness and could lead to a more robust autonomic reaction to orthostatic stress.

Functional connectivity analyses revealed significantly increased thalamic connectivity with bilateral sensorimotor and visuo-occipital cortices in ME/CFS patients at baseline. These regions have previously been implicated in fatigue, sensory overload, and motor slowing in ME/CFS [24], [25], as well as in related central sensitivity syndromes such as fibromyalgia [46], [47]. This increased connectivity, which may reflect impaired filtering, heightened sensory gain, or compensatory over-engagement of thalamocortical circuits, was no longer present at follow-up, suggesting normalization after treatment. Notably, longitudinal within-group analyses in the patient cohort showed post-treatment reductions in the FC of thalamic subcluster for sensorimotor integration [43]. The thalamus, a key relay hub for motor and sensory information [48], [49], has been linked to fatigue across various conditions, including postlilCOVID syndrome [50]. Leitner_et_al. [50] recently reported that postlilCOVID patients with fatigue exhibit aberrant thalamic connectivity with motorlilassociated cortices compared to nonlilfatigued patients, with connectivity strength correlating with fatigue severity and processing speed. While their fatigued cohort exhibited reduced thalamo-motor FC, and our ME/CFS cohort showed thalamic hyperconnectivity, both studies highlight disrupted thalamocortical communication in sensorimotor circuits as a shared neural signature of persistent fatigue. Similar neuroimaging findings in other neurological conditions, including multiple sclerosis and stroke, further implicate thalamocortical circuit dysfunction in fatigue [51], [52], [53], [54].

Further analyses in responders and non-responders showed that normalization of FC in longitudinal analyses was mainly driven by larger FC reductions from pre- to post-treatment in responders, whereas FC changes in non-responders were limited to small clusters. This suggests that clinical improvement in physical functioning in ME/CFS (as measured by a ≥10-point increase in SF-36 Physical Function score) may be mediated by changes in thalamic regulation of cortical networks, particularly those involved in sensorimotor integration. Notably, non-responders showed improvements comparable to responders in processing speed and pain, suggesting that HBOT may exert broader effects beyond physical functioning, potentially involving cognitive and pain-related neural networks that were not captured by our thalamic FC analyses. Importantly, we found no significant volumetric alterations in ME/CFS patients across any comparison, supporting the notion that ME/CFS symptoms may be related to functional network changes rather than macrostructural abnormalities. Taken together, our imaging findings complement the clinical observations and support the hypothesis that disrupted sensorimotor FC, likely driven by dysfunctional thalamic relay, is a feature of ME/CFS, and that HBOT may contribute to rebalancing these circuits.

This study has several limitations. First, the small sample size and observational, non-controlled design limit the generalizability and causal interpretation of our findings. Second, our findings regarding the feasibility and safety of HBOT may not extend to more severely affected, homebound patients, who were excluded from this study. Third, while our imaging results suggest normalization of thalamocortical hyperconnectivity following HBOT, the absence of a sham-treated control group prevents definitive attribution of these changes to the intervention. Furthermore, although our findings imply that functional, rather than structural, brain alterations may underlie key ME/CFS symptoms, future studies should include longitudinal multimodal imaging and consider network dynamics. Finally, several questions remain, including the optimal number and frequency of HBOT sessions, the durability of the reported effects, and the identification of patient subgroups most likely to benefit from HBOT therapy. These aspects will be addressed in our ongoing research, including a second cohort currently undergoing a shortened 20 session protocol. Additionally, laboratory analyses, as well as endothelial function tests, are underway and will be integrated in future analyses.

Strengths of this study include its multidimensional assessment of HBOT effects, integrating patient-reported outcomes with physician assessed functional tests and neuroimaging. This comprehensive approach allowed evaluation of both subjective improvements and objective physiological changes. In particular, the functional brain analyses revealed baseline thalamic hyperconnectivity in ME/CFS patients, which normalized following HBOT and was most pronounced in clinical responders, providing mechanistic plausibility and a potential biomarker of treatment response. The consistent improvements across multiple symptom domains, together with high adherence to the demanding treatment protocol and favorable tolerability, further support the feasibility and clinical relevance of HBOT in this patient population.

## 5. Conclusion

In conclusion, this observational study provides encouraging evidence that HBOT may lead to significant improvements in physical functioning, enhance exercise capacity and muscle strength, improve information processing speed and reduce fatigue and pain in ME/CFS patients. These findings are consistent with results reported in post-COVID patients, despite some variations in treatment protocols. Furthermore, restinglilstate fMRI revealed a post-treatment normalization of thalamocortical hyperconnectivity with sensorimotor and visuoliloccipital networks, with greater reductions associated with more pronounced physical improvement. These results suggest that functional brain network dysregulation, rather than macrostructural abnormalities, may contribute to core ME/CFS symptoms and that such dysregulation may be modulated by HBOT. Given its favorable safety profile, promising clinical outcomes, and high patient adherence, HBOT may represent a potentially valuable therapeutic option for ME/CFS. Larger, randomized controlled trials are needed to confirm its efficacy and to further explore its mechanisms of action.

## Data Availability

The data presented in this study will be available upon request from the corresponding author. Due to the sensitive nature of the data and the ongoing data collection and analysis, the data are not publicly available.

## Contributors

LK oversaw project administration. LK and GC wrote the first draft of the report and performed statistical analysis, prepared the figures of the report and managed electronic data collection. CH helped with electronic data collection. LK conducted the investigation (patient assessments, recruitment and coordination of study appointments) together with CKP, NL, OM, CvH, MK, TH and LR. CKP, KW and RR helped with patient recruitment. MM contributed to the functional connectivity analysis, as well as the reviewing and editing of the imaging analysis sections in this manuscript. SK contributed to the volumetric neuroimaging analysis. CS together with CF did the conceptualization and provided resources and supervision. All authors had full access to all the data in the study and had final responsibility for the decision to submit for publication. GC and LK have accessed and verified the data.

## Institutional Review Board Statement

The Ethics Committee of the Charité - Universitätsmedizin Berlin approved this study in accordance with the 1964 Declaration of Helsinki and its later amendments (protocol code EA1/129/23, date of approval: 04 July 2023).

## Informed Consent Statement

Informed consent was obtained from all subjects involved in the study.

## References

[1] C. Linda, M. C. van Campen, F. W. A. Verheugt, P. C. Rowe, and F. C. Visser, “Cerebral blood flow is reduced in ME/CFS during head-up tilt testing even in the absence of hypotension or tachycardia: A quantitative, controlled study using Doppler echography,” Clin Neurophysiol Pract, vol. 5, pp. 50–58, 2020, doi: 10.1016/j.cnp.2020.01.003.

[2] D. J. Newton, G. Kennedy, K. K. F. Chan, C. C. Lang, J. J. F. Belch, and F. Khan, “Large and small artery endothelial dysfunction in chronic fatigue syndrome,” Int J Cardiol, vol. 154, no. 3, pp. 335–336, Feb. 2012, doi: 10.1016/j.ijcard.2011.10.030.

[3] F. Sotzny et al., “Myalgic Encephalomyelitis/Chronic Fatigue Syndrome – Evidence for an autoimmune disease,” Autoimmun Rev, vol. 17, no. 6, pp. 601–609, Jun. 2018, doi: 10.1016/j.autrev.2018.01.009.

[4] V. A. Spence, F. Khan, G. Kennedy, N. C. Abbot, and J. J. F. Belch, “Acetylcholine mediated vasodilatation in the microcirculation of patients with chronic fatigue syndrome,” Prostaglandins Leukot Essent Fatty Acids, vol. 70, no. 4, pp. 403–407, Apr. 2004, doi: 10.1016/j.plefa.2003.12.016.

[5] J. Hartwig et al., “IgG stimulated β2 adrenergic receptor activation is attenuated in patients with ME/CFS,” Brain Behav Immun Health, vol. 3, p. 100047, Mar. 2020, doi: 10.1016/j.bbih.2020.100047.

[6] J. Słomko et al., “Autonomic Phenotypes in Chronic Fatigue Syndrome (CFS) Are Associated with Illness Severity: A Cluster Analysis,” J Clin Med, vol. 9, no. 8, p. 2531, Aug. 2020, doi: 10.3390/jcm9082531.

[7] Ø. Fluge et al., “Metabolic profiling indicates impaired pyruvate dehydrogenase function in myalgic encephalopathy/chronic fatigue syndrome,” JCI Insight, vol. 1, no. 21, Dec. 2016, doi: 10.1172/jci.insight.89376.

[8] L. Nacul et al., “European Network on Myalgic Encephalomyelitis/Chronic Fatigue Syndrome (EUROMENE): Expert Consensus on the Diagnosis, Service Provision, and Care of People with ME/CFS in Europe,” Medicina (B Aires), vol. 57, no. 5, p. 510, May 2021, doi: 10.3390/medicina57050510.

[9] C. Kedor et al., “A prospective observational study of post-COVID-19 chronic fatigue syndrome following the first pandemic wave in Germany and biomarkers associated with symptom severity,” Nat Commun, vol. 13, no. 1, Dec. 2022, doi: 10.1038/s41467-022-32507-6.

[10] C. Scheibenbogen et al., “Fighting Post-COVID and ME/CFS – development of curative therapies,” Front Med (Lausanne), vol. 10, Jun. 2023, doi: 10.3389/fmed.2023.1194754.

[11] K. A. Seton, J. A. Espejo-Oltra, K. Giménez-Orenga, R. Haagmans, D. J. Ramadan, and J. Mehlsen, “Advancing Research and Treatment: An Overview of Clinical Trials in Myalgic Encephalomyelitis/Chronic Fatigue Syndrome (ME/CFS) and Future Perspectives,” J Clin Med, vol. 13, no. 2, p. 325, Jan. 2024, doi: 10.3390/jcm13020325.

[12] M. A. Ortega et al., “A General Overview on the Hyperbaric Oxygen Therapy: Applications, Mechanisms and Translational Opportunities,” Medicina (B Aires), vol. 57, no. 9, p. 864, Aug. 2021, doi: 10.3390/medicina57090864.

[13] A. Hadanny and S. Efrati, “The Hyperoxic-Hypoxic Paradox,” Biomolecules, vol. 10, no. 6, p. 958, Jun. 2020, doi: 10.3390/biom10060958.

[14] N. Schottlender, I. Gottfried, and U. Ashery, “Hyperbaric Oxygen Treatment: Effects on Mitochondrial Function and Oxidative Stress,” Biomolecules, vol. 11, no. 12, p. 1827, Dec. 2021, doi: 10.3390/biom11121827.

[15] A. A. Katz, S. Wainwright, M. P. Kelly, P. Albert, and R. Byrne, “Hyperbaric oxygen effectively addresses the pathophysiology of long COVID: clinical review,” Front Med (Lausanne), vol. 11, Feb. 2024, doi: 10.3389/fmed.2024.1354088.

[16] F. Liang, N. Kang, P. Li, X. Liu, G. Li, and J. Yang, “Effect of Hyperbaric Oxygen Therapy on Polarization Phenotype of Rat Microglia After Traumatic Brain Injury,” Front Neurol, vol. 12, Jun. 2021, doi: 10.3389/fneur.2021.640816.

[17] S. Akarsu et al., “The efficacy of hyperbaric oxygen therapy in the management of chronic fatigue syndrome,” Undersea Hyperb Med . , vol. 40, no. 2, pp. 197–200, 2013.

[18] E. Van Hoof, D. Coomans, P. De Becker, R. Meeusen, R. Cluydts, and K. De Meirleir, “Hyperbaric Therapy in Chronic Fatigue Syndrome,” J Chronic Fatigue Syndr, vol. 11, no. 3, pp. 37–49, Jan. 2003, doi: 10.1300/J092v11n03_04.

[19] B.-Q. Wu et al., “Effects of Hyperbaric Oxygen Therapy on Long COVID: A Systematic Review,” Life, vol. 14, no. 4, p. 438, Mar. 2024, doi: 10.3390/life14040438.

[20] S. Zilberman-Itskovich et al., “Hyperbaric oxygen therapy improves neurocognitive functions and symptoms of post-COVID condition: randomized controlled trial,” Sci Rep, vol. 12, no. 1, Dec. 2022, doi: 10.1038/s41598-022-15565-0.

[21] M. Catalogna, E. Sasson, A. Hadanny, Y. Parag, S. Zilberman-Itskovich, and S. Efrati, “Effects of hyperbaric oxygen therapy on functional and structural connectivity in post-COVID-19 condition patients: A randomized, sham-controlled trial,” Neuroimage Clin, vol. 36, Jan. 2022, doi: 10.1016/j.nicl.2022.103218.

[22] A. Hadanny et al., “Long term outcomes of hyperbaric oxygen therapy in post covid condition: longitudinal follow-up of a randomized controlled trial,” Sci Rep, vol. 14, no. 1, p. 3604, Feb. 2024, doi: 10.1038/s41598-024-53091-3.

[23] B. Almutairi, C. Langley, E. Crawley, and N. J. Thai, “Using structural and functional MRI as a neuroimaging technique to investigate chronic fatigue syndrome/myalgic encephalopathy: A systematic review,” Aug. 30, 2020, *BMJ Publishing Group*. doi: 10.1136/bmjopen-2019-031672.

[24] J. Boissoneault, J. Letzen, S. Lai, M. E. Robinson, and R. Staud, “Static and dynamic functional connectivity in patients with chronic fatigue syndrome: Use of arterial spin labelling fMRI,” Clin Physiol Funct Imaging, vol. 38, no. 1, pp. 128–137, Jan. 2018, doi: 10.1111/cpf.12393.

[25] J. Boissoneault et al., “Abnormal resting state functional connectivity in patients with chronic fatigue syndrome: An arterial spin-labeling fMRI study,” Magn Reson Imaging, vol. 34, no. 4, pp. 603–608, May 2016, doi: 10.1016/j.mri.2015.12.008.

[26] J. Cotler, C. Holtzman, C. Dudun, and L. A. Jason, “A Brief Questionnaire to Assess Post-Exertional Malaise,” Diagnostics, vol. 8, no. 3, p. 66, Sep. 2018, doi: 10.3390/diagnostics8030066.

[27] B. M. Carruthers et al., “Myalgic encephalomyelitis: International Consensus Criteria,” J Intern Med, vol. 270, no. 4, pp. 327–338, Oct. 2011, doi: 10.1111/j.1365-2796.2011.02428.x.

[28] L. Bateman et al., “Myalgic Encephalomyelitis/Chronic Fatigue Syndrome: Essentials of Diagnosis and Management,” Mayo Clin Proc, vol. 96, no. 11, pp. 2861–2878, Nov. 2021, doi: 10.1016/j.mayocp.2021.07.004.

[29] J. E. Ware, “SF-36 Health Survey Update,” Spine (Phila Pa 1976), vol. 25, no. 24, pp. 3130–3139, Dec. 2000, doi: 10.1097/00007632-200012150-00008.

[30] A. Brigden, R. M. Parslow, D. Gaunt, S. M. Collin, A. Jones, and E. Crawley, “Defining the minimally clinically important difference of the SF-36 physical function subscale for paediatric CFS/ME: triangulation using three different methods,” Health Qual Life Outcomes, vol. 16, no. 1, p. 202, Oct. 2018, doi: 10.1186/s12955-018-1028-2.

[31] T. Chalder et al., “Development of a fatigue scale,” J Psychosom Res, vol. 37, no. 2, pp. 147–153, Feb. 1993, doi: 10.1016/0022-3999(93)90081-P.

[32] D. S. Bell, The doctor’s guide to chronic fatigue syndrome: understanding, treating, and living with CFIDS. Addison-Wesley Pub. Co, 1994.

[33] B. Jäkel et al., “Hand grip strength and fatigability: correlation with clinical parameters and diagnostic suitability in ME/CFS,” J Transl Med, vol. 19, no. 1, p. 159, Dec. 2021, doi: 10.1186/s12967-021-02774-w.

[34] S. Vernino et al., “Postural orthostatic tachycardia syndrome (POTS): State of the science and clinical care from a 2019 National Institutes of Health Expert Consensus Meeting - Part 1,” Autonomic Neuroscience, vol. 235, p. 102828, Nov. 2021, doi: 10.1016/j.autneu.2021.102828.

[35] A. Smith, “Symbol Digit Modalities Test,” Jun. 13, 2016. doi: 10.1037/t27513-000.

[36] R. W. Bohannon and R. Crouch, “1-Minute Sit-to-Stand Test,” J Cardiopulm Rehabil Prev, vol. 39, no. 1, pp. 2–8, Jan. 2019, doi: 10.1097/HCR.0000000000000336.

[37] P. A. Harris et al., “The REDCap consortium: Building an international community of software platform partners,” J Biomed Inform, vol. 95, p. 103208, Jul. 2019, doi: 10.1016/j.jbi.2019.103208.

[38] B. Fischl et al., “Whole Brain Segmentation,” Neuron, vol. 33, no. 3, pp. 341–355, 2002, doi: 10.1016/S0896-6273(02)00569-X.

[39] O. Esteban et al., “fMRIPrep: a robust preprocessing pipeline for functional MRI,” Nat Methods, vol. 16, no. 1, pp. 111–116, Jan. 2019, doi: 10.1038/s41592-018-0235-4.

[40] S. Whitfield-Gabrieli and A. Nieto-Castanon, “*Conn*: A Functional Connectivity Toolbox for Correlated and Anticorrelated Brain Networks,” Brain Connect, vol. 2, no. 3, pp. 125–141, Jun. 2012, doi: 10.1089/brain.2012.0073.

[41] Y. Behzadi, K. Restom, J. Liau, and T. T. Liu, “A component based noise correction method (CompCor) for BOLD and perfusion based fMRI,” Neuroimage, vol. 37, no. 1, pp. 90–101, Aug. 2007, doi: 10.1016/j.neuroimage.2007.04.042.

[42] R. S. Desikan et al., “An automated labeling system for subdividing the human cerebral cortex on MRI scans into gyral based regions of interest,” Neuroimage, vol. 31, no. 3, pp. 968–980, Jul. 2006, doi: 10.1016/j.neuroimage.2006.01.021.

[43] O. J. Boeken, E. C. Cieslik, R. Langner, and S. Markett, “Characterizing functional modules in the human thalamus: coactivation-based parcellation and systems-level functional decoding,” Nov. 01, 2023, *Springer Science and Business Media Deutschland GmbH*. doi: 10.1007/s00429-022-02603-w.

[44] T. Robbins et al., “Hyperbaric oxygen therapy for the treatment of long COVID: early evaluation of a highly promising intervention,” Clinical Medicine, vol. 21, no. 6, pp. e629–e632, Nov. 2021, doi: 10.7861/clinmed.2021-0462.

[45] J. Lindenmann et al., “Immediate and Long-Term Effects of Hyperbaric Oxygenation in Patients with Long COVID-19 Syndrome Using SF-36 Survey and VAS Score: A Clinical Pilot Study,” J Clin Med, vol. 12, no. 19, p. 6253, Sep. 2023, doi: 10.3390/jcm12196253.

[46] P. Flodin, S. Martinsen, M. Löfgren, I. Bileviciute-Ljungar, E. Kosek, and P. Fransson, “Fibromyalgia Is Associated with Decreased Connectivity Between Pain- and Sensorimotor Brain Areas,” Brain Connect, vol. 4, no. 8, pp. 587–594, Oct. 2014, doi: 10.1089/brain.2014.0274.

[47] D. J. Kim, M. Lim, J. S. Kim, and C. K. Chung, “Structural and functional thalamocortical connectivity study in female fibromyalgia,” Sci Rep, vol. 11, no. 1, p. 23323, Dec. 2021, doi: 10.1038/s41598-021-02616-1.

[48] R. W. Guillery and S. M. Sherman, “Thalamic Relay Functions and Their Role in Corticocortical Communication,” Neuron, vol. 33, no. 2, pp. 163–175, Jan. 2002, doi: 10.1016/S0896-6273(01)00582-7.

[49] S. Murray Sherman, “Chapter 4 Thalamic relay functions,” 2001, pp. 51–69. doi: 10.1016/S0079-6123(01)34005-0.

[50] M. Leitner et al., “Changes in thalamic functional connectivity in post-Covid patients with and without fatigue,” Neuroimage, vol. 301, p. 120888, Nov. 2024, doi: 10.1016/J.NEUROIMAGE.2024.120888.

[51] F. Capone, S. Collorone, R. Cortese, V. Di Lazzaro, and M. Moccia, “Fatigue in multiple sclerosis: The role of thalamus,” Jan. 01, 2020, *SAGE Publications Ltd*. doi: 10.1177/1352458519851247.

[52] M. Hidalgo de la Cruz et al., “Abnormal functional connectivity of thalamic sub-regions contributes to fatigue in multiple sclerosis,” Multiple Sclerosis Journal, vol. 24, no. 9, pp. 1183–1195, Aug. 2018, doi: 10.1177/1352458517717807.

[53] J. Wang et al., “Functional connectome hierarchy of thalamus impacts fatigue in acute stroke patients,” Cerebral Cortex, vol. 34, no. 2, Feb. 2024, doi: 10.1093/cercor/bhad534.

[54] C. Finke et al., “Altered basal ganglia functional connectivity in multiple sclerosis patients with fatigue,” Multiple Sclerosis Journal, vol. 21, no. 7, pp. 925–934, Jun. 2015, doi: 10.1177/1352458514555784.

